# The Syphilis Point of Care Rapid Testing and Immediate Treatment Evaluation (SPRITE) Study

**DOI:** 10.1101/2024.05.24.24307902

**Authors:** Lucy Mackrell, Megan Carter, Maggie Hoover, Patrick O’Byrne, Natasha Larkin, Felicia Magpantay, Sicheng Zhao, Bradley Stoner, Melissa Richard-Greenblatt, Kira Mandryk, Kandace Belanger, Jennifer Burbidge, Gilles Charette, Gabrielle Deschenes, Duy Dinh, Amanda Featherstone, Farhan Khandakar, Jorge Martinez-Cajas, Vanessa Tran, Nicole Szumlanski, Stephanie Vance, Sahar Saeed, the SPRITE Study

## Abstract

**Background:** In the last decade, the rates of infectious syphilis have increased by 505% in Ontario, Canada. Underserved populations–people who use drugs, un(der)housed individuals, and those living in rural and remote areas - face unique social and healthcare challenges that increase their vulnerability to syphilis infections and hinder their access to timely diagnosis and treatment. Given the shift in epidemiology and the geographic disparities in resources, there is no one-size-fits-all solution to this complex issue. Access to low-barrier diagnostics, such as point-of-care (POC) tests for HIV and hepatitis C, has been shown to be an effective strategy for reaching underserved people outside of traditional healthcare settings. However, there is a paucity of evidence regarding the effectiveness of POC tests for other sexually transmitted and blood-borne infections, particularly in non-urban, rural, and remote settings.

**Methods/Design:** The Syphilis Rapid Point-of-Care Testing and Immediate Treatment Evaluation (SPRITE) Study includes nine Public Health Units (PHUs) in Ontario. The INSTI® Multiplex HIV-1/HIV-2/Syphilis Antibody Test will be used to evaluate a “rapid test and immediate treatment” outreach model of care targeting people who are un(der)housed or who use drugs at multiple community-based settings anchored to their respective PHU. Implementation of the model of care will be evaluated using the Reach, Effectiveness, Adoption, Implementation, and Maintenance (RE-AIM) Framework, following a community-based participatory approach. Empirical results will inform network models to estimate the population-level impact of using POC test to curb transmission.

**Discussion:** Urgent, tailored, and equitable action is needed to address the alarming rise in syphilis rates in Canada. This study assesses the real-world effectiveness of syphilis POC tests in breaking barriers and bringing services to the population at the highest risk. Results will inform future implementation, build capacity, and provide the evidence necessary for program decision-making.

**STRENGTHS AND LIMITATIONS OF THIS STUDY:**

- This study employs a comprehensive evaluation framework, utilizing various approaches and tools encompassing quantitative and qualitative methods to address our research questions. This multiplicity enables enhanced triangulation of research findings.
- Including various public health units addresses a gap in research by acknowledging and investigating the unique challenges faced by underserved communities living in diverse non-urban, rural, and remote settings.
- Leveraging academic, public health, and community partnerships enhances this study’s potential for real-world impact by incorporating diverse perspectives, expertise, and collaborative efforts.

## INTRODUCTION

Syphilis is a treatable sexually transmitted and blood-borne infection (STBBI) caused by the bacterium *Treponema pallidum pallidum(1)*. A one-time intramuscular injection of 2.4 million units of benzathine penicillin G can effectively treat early infections (< 1-year duration), preventing a myriad of serious health consequences (1, 2). Syphilis is described as having several predictable stages (Figure 1). While not every individual goes through each stage, the typical trajectory is as follows (3). The primary phase, typified by replicating spirochetes at the inoculation site, induces a local inflammatory response, giving rise to one or more chancres around three weeks (9-90 days) following infection. Secondary syphilis typically occurs 4 to 10 weeks after the chancre because of spirochete dissemination. At this stage, any organ can be affected; however, mucocutaneous lesions are predominant, most seen as a maculopapular rash on the palms and soles. These symptoms may last anywhere from 3-12 weeks, and occasional relapses may be seen within the first few years in persons who have not received treatment. A period in which individuals show no clinical signs or symptoms of primary or secondary syphilis follows secondary syphilis. This period is divided into ‘early non-primary non-secondary’ (early latent) (< one year) and ‘late’ (late latent) (> one year). Without treatment within 5-30 years, up to 35% of individuals with late syphilis develop clinical manifestations (tertiary syphilis), including neurosyphilis, cardiovascular syphilis, or gummatous syphilis. (3). Neurosyphilis, ocular syphilis, and otosyphilis can occur at any stage (4). Most syphilis transmission occurs through sexual contact and is the most infectious during the early stages (primary, secondary, and early non-primary non-secondary). The next most common form of transmission is across the placenta (2), which results in congenital syphilis (2). Congenital syphilis is treatable in utero, but without treatment, it can have devastating effects, including stillbirth, neonatal death, or severe health effects such as bone damage or nerve problems (3, 5). In rare instances, when there are maternal genital lesions, syphilis can be transmitted at birth through the infant’s contact with the birth canal (6).

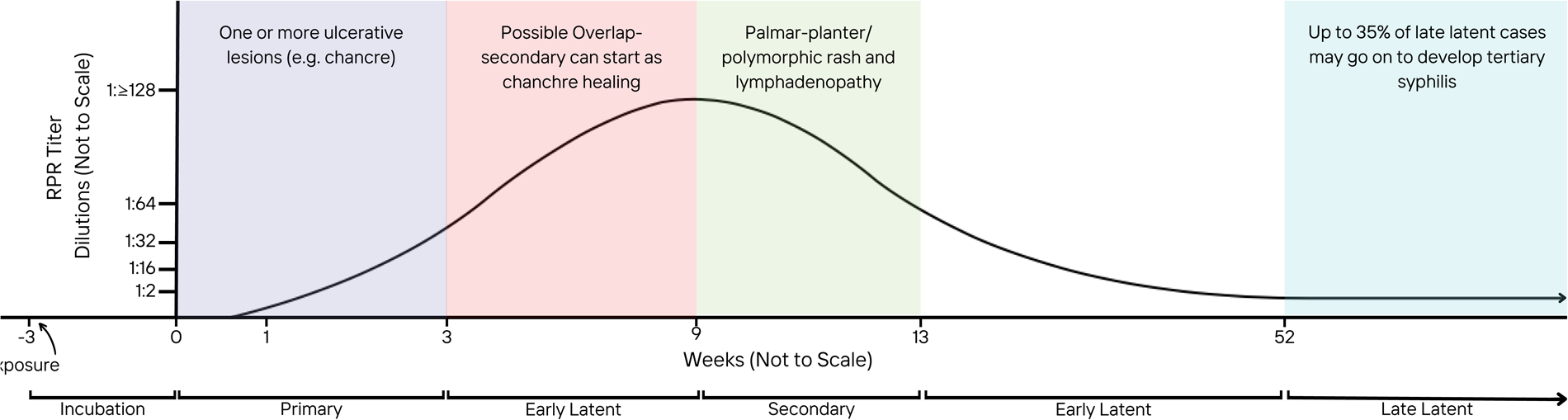

## Epidemiology

Canada has seen a marked increase in infectious syphilis, with a 109% increase in cases between 2018 and 2022, and geographic heterogeneity across the country (7). In Ontario, Canada’s most populous province, between 2013 and 2022, infectious syphilis rates grew from 3.9 to 23.6 cases per 100,000 people (8, 9). This marked surge was paired with a significant demographic shift. Historically, males (mainly men who have sex with men (MSM)) were disproportionately affected by syphilis. However, between 2013 and 2022, females of reproductive age (aged 15 to 39) were identified as the fastest-growing at-risk population (8). As a result, cases of congenital syphilis soared by 2600%, with 27 cases reported in 2022 (8, 10). Within Ontario, Public Health Units (PHUs) serving non-urban areas are experiencing the greatest rate increase (11). In December 2022, Kingston, Frontenac and Lennox & Addington Public Health (KFL&A PH) declared a syphilis outbreak as their rates of infectious syphilis (41.2 per 100,000) surpassed the provincial average (23.6 per 100,000), and five congenital cases (2.8 cases per 1000 live births) were reported (8).

Underserved communities including people who use drugs, and un(der)housed individuals are known to be disproportionately affected by syphilis (8, 12, 13). In 2022, the age-standardized rate of infectious syphilis among females 15 to 44 years of age was almost three times higher in neighbourhoods with the most materially related marginalization, compared to those with the least materially related marginalization (8). Housing instability and stigmatization syndemic to substance misuse, along with co-occurring comorbidities and complex mental health needs, increase barriers to accessing to health care. Barriers to care, whether due to competing survival needs or discrimination within the health care system, delays diagnosis and treatment (1, 7, 10, 14–17). This syndemic is exacerbated in rural and remote communities due to resource limitations (18).

### Diagnosis

Diagnostic screening for syphilis in Ontario is typically conducted within a healthcare setting based on clinical guidelines outlined by PHO and the Canadian Guidelines on Sexually Transmitted Infections (19, 20). Syphilis serology testing is indicated for the diagnosis of suspected syphilis infections and post-treatment monitoring. Screening is recommended for anyone sexually active, having a new partner, or upon request, as well as pregnant individuals during their first trimester or first prenatal visit. Additional screening at mid-gestion/third trimester (28-32 weeks) and delivery vary as either universal or risk-based recommendations (19, 20). PHO follows a reverse algorithm, starting with a chemiluminescent microparticle immunoassay (CMIA), a qualitative immunoassay that detects treponemal antibodies (IgG and IgM). If reactive, confirmatory rapid plasma reagin (RPR) is performed. Any non-reactive confirmatory RPRs will have the *Treponema pallidum* particle agglutination (TPPA) assay performed to aid in diagnosing current/past syphilis infections. Turnaround time for syphilis results is up to 3 business days from receipt at PHO’s laboratory for non-reactive specimens and up to 6 business days for reactive specimens (19).

The current model of care is inadequate to address infectious syphilis in underserved populations, as social and economic challenges increase the likelihood of becoming infected and impact timely testing and treatment (1, 7, 10, 15, 16, 21). Low-barrier interventions using point-of-care (POC) tests are acceptable and reliable for other STBBIs (22–28). On March 27^th^, 2023, Health Canada approved – INSTI® Multiplex HIV-1/2 Syphilis Antibody Assay – an all-in-one rapid POC test which detects syphilis and HIV antibodies in under five minutes (29). This approval marked a significant advancement in reducing the spread of STBBIs in Canada. However, innovations in technology only hold promise to improve health outcomes if paired with effective implementation in real-world practice. While the INSTI® Multiplex POC product monograph reports 95-100% sensitivity (30), more real-world studies are needed to examine accuracy in a community setting and in populations with low RPR concentrations (<1:8 dilutions) (Figure 1). Expanding POC tests to non-healthcare settings is a crucial strategy to reach undiagnosed individuals, but few studies have evaluated the effectiveness and feasibility of this approach in various settings (31–33). Here we describe the methodology of the Syphilis Rapid Point-of-Care Testing and Immediate Treatment Evaluation (SPRITE) Study, wherein the overall goal is to evaluate the implementation and effectiveness of a community-based outreach model of care aimed at reducing barriers to timely testing and treatment of syphilis across PHUs in Ontario.

## METHODS AND ANALYSIS

### Setting

Ontario Public Health Units (PHUs): Kingston, Frontenac and Lennox & Addington Public Health (KFL&APH), Hastings Prince Edward Public Health (HPEPH), Leeds Grenville & Lanark District Health Unit (LGLDHU), Thunder Bay District Health Unit (TBDHU), Ottawa Public Health (OPH), Algoma Public Health (APH), Renfrew County and District Health Unit (RCDHU), Middlesex-London Health Unit (MLHU), and Porcupine Health Unit (Porcupine HU). HPEPH (population of 171,450), KFL&APH (206,962), APH (112,764), TBDHU (152,885), and Porcupine HU (81,188) belong to Peer Group C (i.e., sparsely populated mix of rural and urban), while LGLDHU (179,830) and RCDHU (107,522) are considered more rural (Peer Group D), and OPHU (1,017,889) and MLHU (500,563) more urban (Peer Group B) (34). The study is open to expanding to additional public health units serving Ontario’s non-urban, rural, and remote populations.

### Model of Care

The SPRITE Study uses the INSTI® Multiplex HIV-1/HIV-2/Syphilis Antibody Test to evaluate a “rapid test and immediate treatment” outreach model of care, bringing services to the population at the highest risk through public health outreach services. Various outreach models, including organized events at community-based organizations (i.e. supervised consumption sites, routine harm reduction outreach (at a specific location or with the use of mobile units), or un(der)housed outreach (i.e. shelter visits) based on their capacity to test and treat.

As per medical directives, trained nurses will use the INSTI® POC test to screen for syphilis and HIV infections. The POC test comprises individually packaged test kits with a lancet, capillary fill pipette, alcohol swab, membrane unit, sample diluent, colour developer, and clarifying solution, and as directed, must be administered by trained healthcare providers (HCPs). Blood from a finger prick is collected via lancet and capillary fill pipette, mixed using sample diluent, added to the membrane unit, followed individually by the colour developer and lastly, clarifying solution. Results are read after the addition of the three vials is complete (∼one minute) and not after five minutes following the addition of the clarifying solution. A reactive syphilis POC test will be considered a suspect case (30). Participating PHUs have each developed a medical directive allowing trained outreach nurses to immediately treat these suspect syphilis cases with a single dose of benzathine penicillin G (2.4 million units via intramuscular injection).

Participants who screen reactive on POC test for HIV will be linked to HIV care. Regardless of POC test result, clients will receive STBBI education and will be asked to provide a single venous blood specimen (serum) for confirmatory serologic testing – to confirm POC test results and aid in determining future treatment response. After confirmatory serologic results are received by the PHU from the PHO laboratory, a nurse initiates follow-up with the participant if the serologic results are reactive, especially if the response results differ from the POC test result. Determining adequate treatment depends on the infection’s staging, which is based on confirmatory serologic results, timing of past testing and results, and clinical judgment. As per each PHU medical directive, the public health nurse collaborates with public health physicians to determine staging. If further treatment is required based on staging, the public health nurse will offer this to the patient. Interval timing for post-treatment testing is based on the staging of the syphilis infection and will assess adequate treatment response.

### Data Collection

In addition to collecting routine data as part of the public health intake forms, ie. demographics, sexual preferences and practices, and risk factors (e.g. injection drug use, sex work), details of the intervention (i.e. location, incentives), implementation outcomes, and testing/treatment results are also collected (see *Table 1*). Outreach nurses at each PHU will extract this data using standardized case report forms from their client records (electronic medical records or paper charts) and all serology results will be obtained from the PHO Laboratory. All research data will be entered into a centralized and standardized electronic database via Medallia.

**Table 1:** Variables being collected through outreach.

| **Variable Type** | **Description / Guidance** |
| --- | --- |
| **Intervention date** | Date person was assessed during outreach |
| **PHU** | Name of PHU where person was tested |
| **Location- Event** | Location or event where person was tested, multiple choice:<br>POC test Blitz/Event; General Outreach; Community service hub<br>(i.e. Consumption center, group programming, etc.); Congregate<br>Setting/Shelter; Public Health Clinic; Primary Care Centre |
| **Demographics** | Sex, Age (year of birth) |
| **MDI/SDI** | Postal Code linked to material/social deprivation index (35) |
| **Community services** | Is the person already connected with other community services? |
| **Risk Factors** | Person works in the sex trade (past/present)<br>Person use and type of injection drugs<br>Person use and type of inhalation drugs<br>Person has sex with anonymous partners<br>Person identifies as gay, bisexual, or other man who has sex with men<br>Person is underhoused or homeless<br>Person is pregnant or breastfeeding |
| **Number of partners** | The number of sexual partners the person had in the last 6 months |
| **Symptoms Present** | Does the person have symptoms for HIV or syphilis? |
| **Previously Tested for STBBIs** | Has the person been previously tested for STBBIs? Date of last STBBI testing |
| **Previously Tested for Syphilis** | Has the person been previously tested for syphilis? Date |
| **History of Syphilis** | Has the person had a previous syphilis diagnosis? If yes, was it treated? |
| **History of HIV** | Has the person received a diagnosis of HIV? If yes, are they on treatment? Connected to care? |
| **POC Test Completed** | Was a POC test completed for this person? |
| **Multiple POC Tests Required** | Did this result require more than one POC test (e.g. previous tests were not performed correctly/invalid)? |
| **Incentive** | Was an incentive provided? |
| **Reason for No Incentive** | If no incentive provided, why not? |
| **Serology Completed** | Was confirmatory serology completed? |
| **Syphilis POC Test Result** | What was the result of POC test for syphilis? |
| **Syphilis Treat (POC Test)** | Was the person treated at POC based on POC test syphilis results? |
| **Syphilis Serology Result** | What was the serology result for syphilis? |
| **Positive Syphilis Serology- Type** | From reactive syphilis serology, is this a previous positive or new positive? (include new and reinfections as new positives) |
| **Syphilis Serology RPR** | From reactive syphilis serology results, enter the RPR if available. |
| **HIV POC Test Result** | What was the result of POC test for HIV? |
| **HIV Serology Results** | What was the serology result for HIV? |
| **Positive HIV Serology-Type** | From reactive HIV serology, is this a previous positive or new positive? |

### Objective 1: Evaluate the real-world effectiveness and implementation of syphilis “POC Test and Treat” model of care using the RE-AIM framework

#### RE-AIM Framework

We will use the Reach, Effectiveness, Adoption, Implementation, and Maintenance (RE-AIM) framework. Under the implementation science umbrella, the RE-AIM model provides a systematic and comprehensive approach to evaluating the implementation and impact of health interventions (36, 37). RE-AIM was created to improve the translation of scientific advances into practice and is one of the most used approaches for policy and program evaluation in public health research (38). Through this study, we will assess the reach, effectiveness, adoption, and implementation of the SPRITE study.

#### Reach

Objective 1a) Assess the extent to which the intervention reaches the target population, including examining the why and how of who is, and is not, being reached (36).

*Methods:* The demographic data on participating in the intervention will be used to evaluate if the target population is being reached by comparing this to historical population-level testing data. This study will also assess individual-level decision-making around POC test and syphilis testing more generally. To this end, we will use qualitative methods to conduct individual interviews and utilize grounded theory methodology to identify factors related to accessing syphilis testing for our target population (39).

*Sampling:* We will initially conduct purposive sampling to identify and conduct semi-structured interviews with two to three members of the target population being offered the intervention. We will use theoretical sampling from the initial themes identified in these interviews, which considers the preliminary data and gaps when selecting future participants. These rounds of analysis and theoretical sampling will continue until theoretical saturation is reached, meaning the point where no additional themes or insights emerge from the data (approximately 10-20 individuals).

*Analysis:* Memoing, where researchers write down ideas about evolving theory to discover patterns, will occur throughout the research process (40). An analytic direction will be developed through the initial coding of interview transcripts in NVivo, followed by focused coding, whereby initial codes are synthesized into categories to explain larger data segments. By looking at how the categories and memos fit best together, we will map the relationship between them through coherent analysis, eventually identifying recurring themes that assist in offering an explanation of syphilis testing among the target population (39).

#### Effectiveness

Objective 1bi) Determine the diagnostic accuracy of INSTI® in non-clinical settings (41).

*Methods:* Diagnostic test accuracy will be measured against serological results. Sensitivity will be measured as the proportion of true positive POC tests of all participants with syphilis.

Specificity is the percentage of true negative POC tests out of all subjects who do not have syphilis. Positive Predictive Value will be calculated based on the number of true positives out of all the positive POC test findings. Negative Predictive Value is the number of true negatives out of all the negative POC test findings.

*Sample Size*: The manufacturer estimates sensitivity between 95% and 100%(30). The sample size of 346 was calculated based on detecting a sensitivity of 90% if the prevalence of syphilis is 10% in the study population, with a 10% margin of error and 95% confidence. The SPRITE study targets a total sample size of 980 paired POC tests and serological tests to allow for lower real-world sensitivity and stratify the analysis by PHUs and by rapid plasma regain titre (non-reactive, <1:8, ≥1:8 dilutions) if sample size in each stratum allows.

Objective 1bii) Estimate the effectiveness of the POC test and treatment model of care reaching undiagnosed (infectious) individuals and minimizing the time between diagnosis and treatment.

*Methods:* We will compare the median time between diagnosis and treatment, loss to follow-up, and test positivity rates before and after the intervention by PHU using an interrupted time series design, using a segmented linear mixed effects model (42). This regression-based method expands upon a basic pre-post comparison design by modeling outcome trends as a function of time (slopes) and using PHUs who do not initiate the intervention as control groups. The attributable impact of the intervention is measured as the difference between the estimated model representing the post-treatment slope derived from observed data and the counterfactual slope (an extension of the pre-treatment slope), along with any immediate level change (43).

#### Adoption

Objective 1c) Assess the willingness and capacity of PHUs and community organizations to adopt the intervention.

Assessing the feasibility of POC test and treatment provision largely borrows from a framework and set of questions developed by the WHO ProSPeRo Network (44), with some adaptations (See *Table 2*). Public health leads participating in the SPRITE study will ask staff or partners participating in the study to complete the adapted WHO feasibility survey online using Medallia two to three months after implementation, ideally after a larger-scale testing event.

The adapted survey contains a series of Likert items consisting of a discrete number of response choices per question. Staff will be asked to select a level of difficulty corresponding to tasks and rank their level of agreement with different statements related to the POC test and treatment intervention, resulting in subdomain scores in the areas of learnability, willingness, suitability, and satisfaction (Table 2). Responses to Likert questions will be scored on a scale of 1 to 5, with 1 being the least favourable response and 5 the most. Each participant’s subdomain score will be calculated by taking the mean of all the subdomain question scores (excluding any ‘N/A’ responses) using a summated scores method, where the same weight was considered for all questions in each subdomain.

The survey will also have open-text boxes for each domain to allow for further comment. The survey will be pre-tested with the working group and updated before deployment.

To understand willingness and capacity in different PHUs, a qualitative content analysis of documents and open-ended text from the surveys will be conducted. Based on the information gleaned from surveys, focus groups and/or individual interviews will be held with intervention agents (medical officers of health, managers, administrators, outreach nurses, harm reduction workers) from each participating PHU and community-based organization (CBO) (n=∼30) to clarify and expand upon themes identified in the surveys. Interviews will focus on identifying and overcoming logistical and organizational barriers (e.g., time, access, training) as well as systemic and structural barriers (e.g., racism, stigma, marginalization) which could impede successful outreach, testing, and treatment. Using descriptive and thematic analysis, focus group and interview transcripts will be systematically coded and analyzed to expand upon what was learned in the survey and identify any new themes that emerged (45).

**Table 2:**
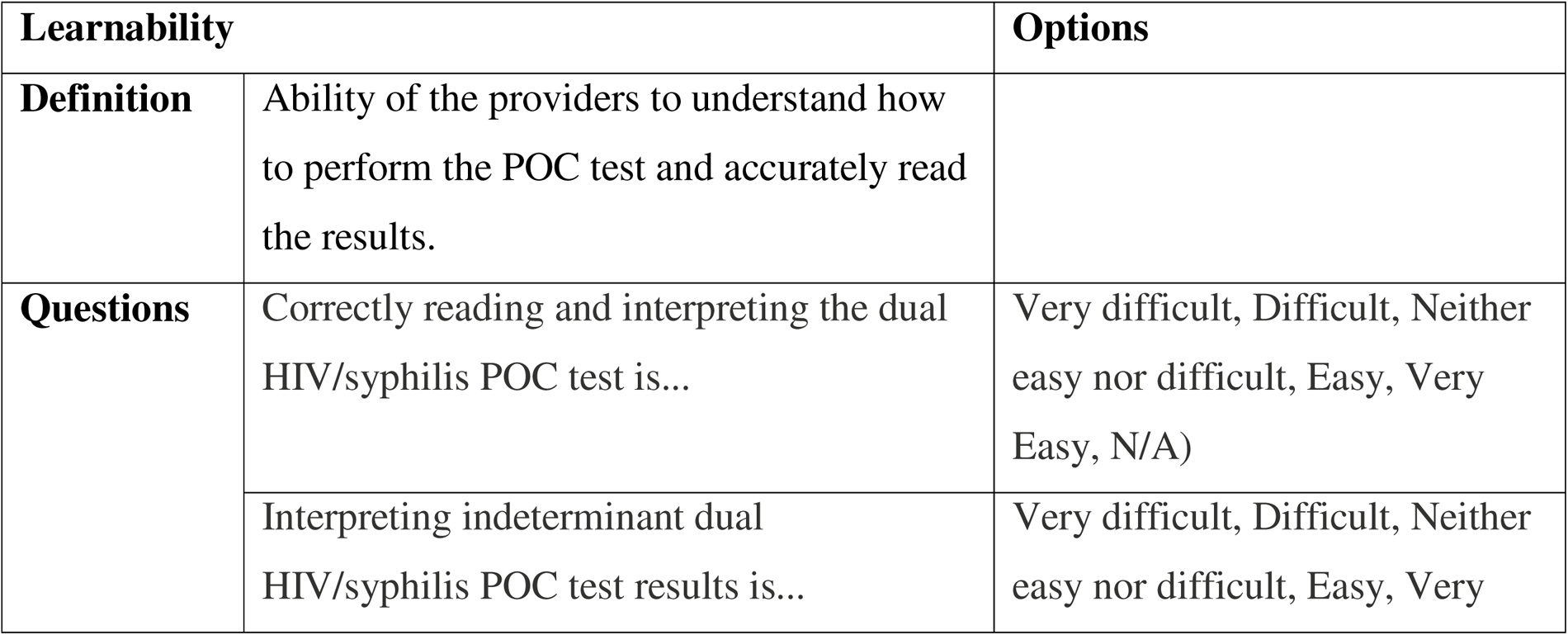
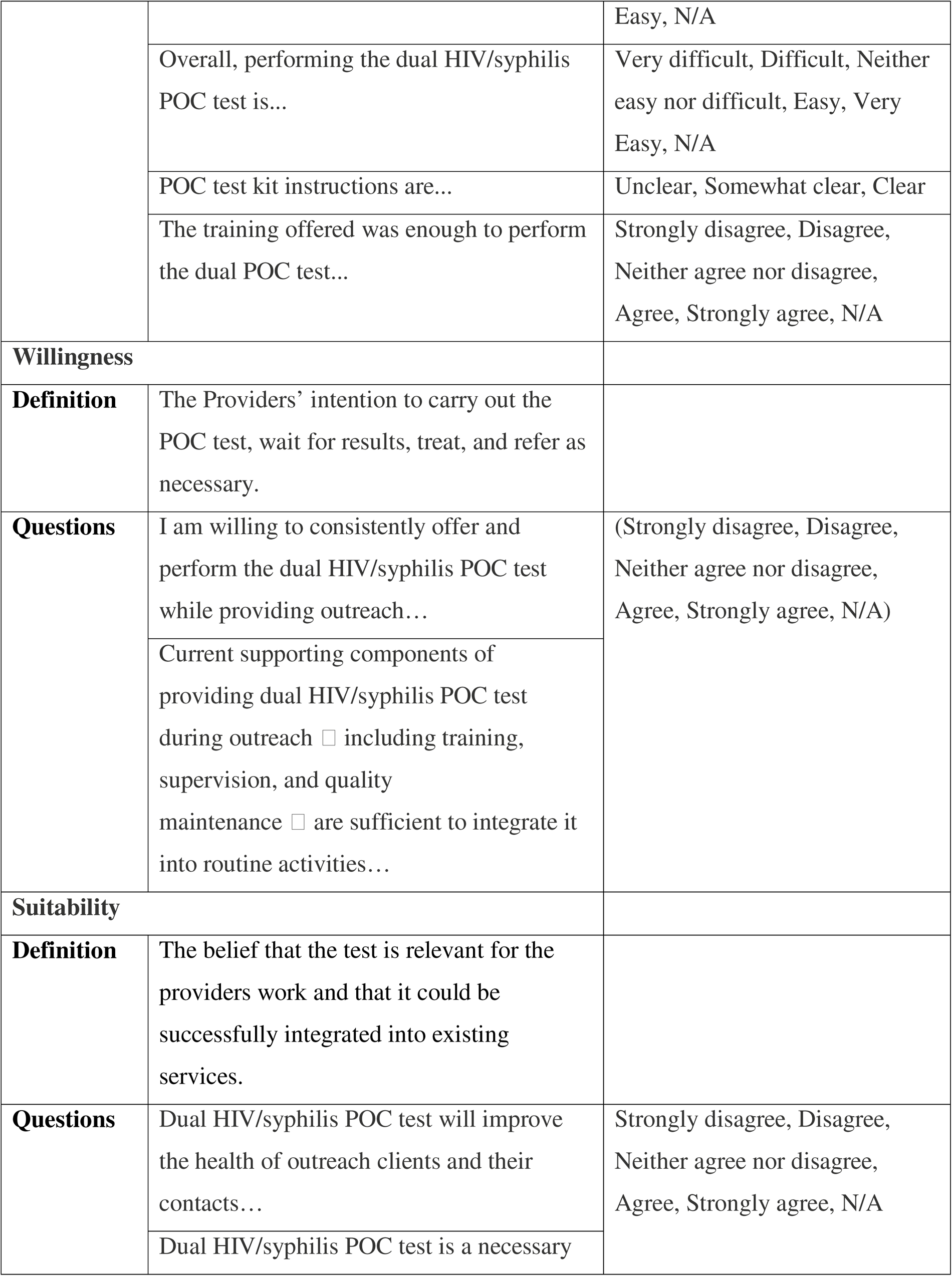
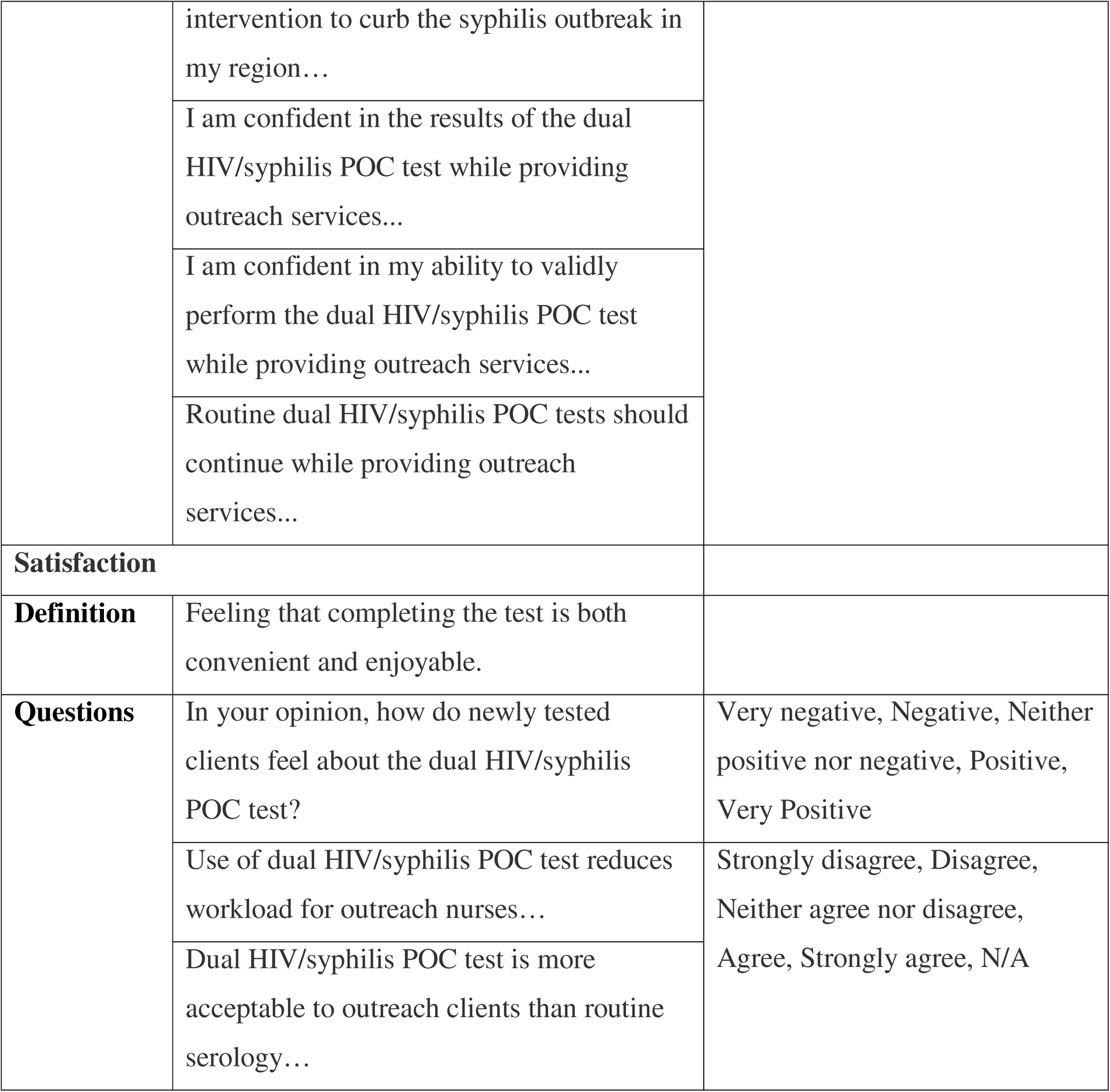
Healthcare Provider (HCP) POC Test Implementer Survey - subdomain definitions and related questions.

#### Implementation

Objective 1d) Understand the modifications made to the intervention during **implementation** and the rationale for adapting the protocol to different settings and contexts (36).

We will document differences in implementation, time-to-implementation, barriers, facilitators, and lessons learned across PHUs. This will include understanding regional context and how that may affect implementation. The number and type of health care providers delivering POC testing and treatment, the number and types of incentives given to individuals being tested, the number and type of outreach activities planned and delivered, medical directives and program policies and procedures related to the provision of POC test and treat, and cost of providing POC test will be tracked over time in each PHU.

### Objective 2: Estimate the potential impact of the POC test and treat model of care using network models

Mathematical modelling will be used to estimate the population-level impact of this test-and-treat model of care on ongoing transmission. The proposed model will provide insights into the impact of deploying intervention variations in terms of the number of potential cases averted relative to the current model of care.

*Data:* Most intervention parameters will be estimated using this primary data from objective 1. Public health surveillance data will be used to obtain 1) demographics (size of populations, age structure) 2) biological parameters (infection rates, stage, progression rate); 3) behavioural data (sexual contacts, testing behaviour) and a systematic review of the literature will be used to collect additional parameters as needed.

*Analysis*: We will use a novel network model to project syphilis transmission. This approach extends from a well-known compartmental susceptible-infectious-recovery model ((Figure 2A) to incorporate a finite set of contacts to whom transmission can occur. Together, this forms a “mixing complex network,” which can be visualized as clusters of nodes and links (Figure 2B)(46). Network modelling requires a more complex theoretical and computational analysis and is more appropriate for STBBI. As illustrated in Figure 2C, the choice of model can lead to stark differences as these two models were calibrated to have the same basic reproduction number (R_0_), but the standard SIR model forecasts a worse epidemic than a network model. The “*EpiNetPerco*” R package will compile different percolation-based methods (network analysis) for disease transmission networks (47–49). Using this package, we will be able to study how quickly the POC test and treat model of care can stop transmission within underserved populations to impact a broader sexually active community. The model will be calibrated with data on sexual behaviours, the annual number of syphilis diagnoses, and the annual number of individuals screened and treated, accounting for observed diagnostic and treatment delays from 2020-2023(50). We will then compare the predicted number of incident syphilis cases with our intervention to the incidence that would have been observed had our study not been implemented over both medium-and long-term time horizons.

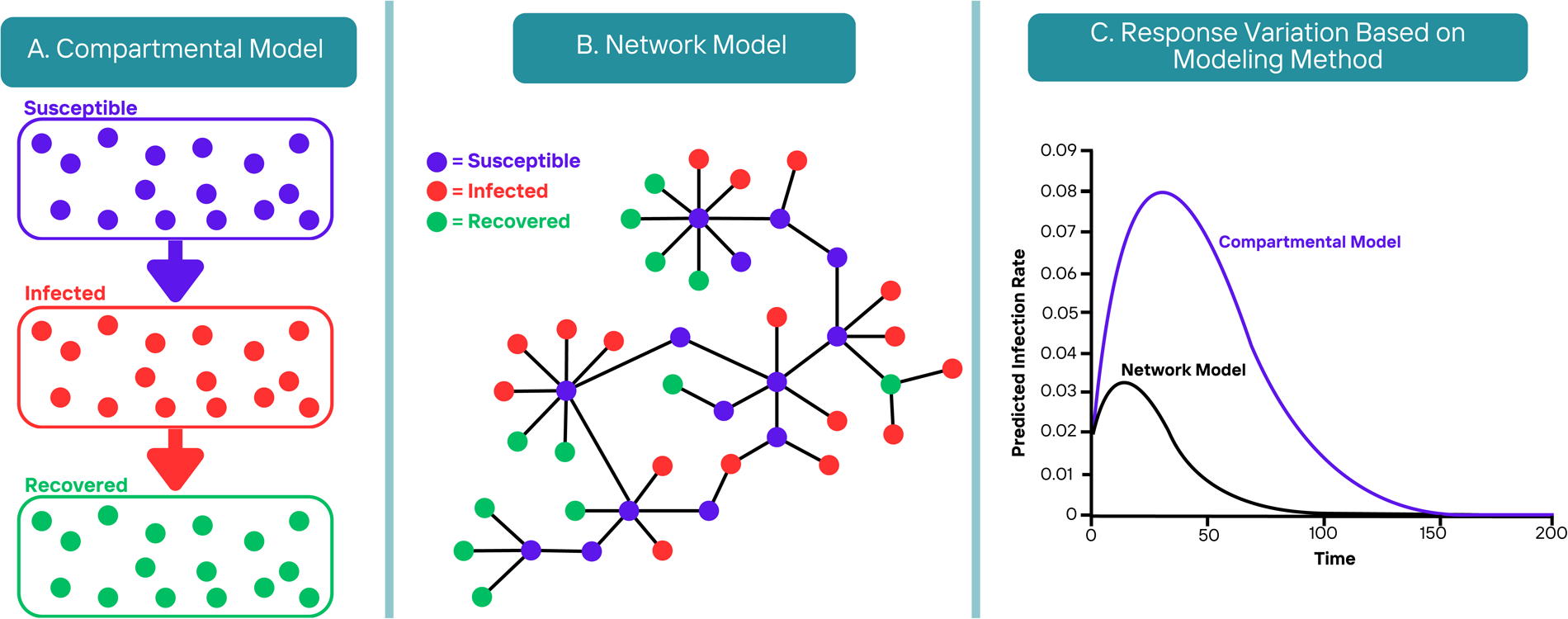

### Objective 3: Create a real-time exchange of information between communities, PHUs, and other relevant knowledge users

Using the Community-Based Participatory Research framework, knowledge users (KU) are engaged and integrated in knowledge translation (51, 52).

*Methods*: Meaningful partnerships with trusted CBOs that serve underserved communities are integral to this research due in part to the hidden nature of this vulnerable group (52). This study leverages relationships between PHUs and CBOs to build a network (Figure 3), facilitating a real-time exchange of information between communities, PHUs, and other relevant knowledge users. Over the last year, the research team from Queen’s University has regularly met with a syphilis steering committee representing clinicians, outreach nurses, and the sexual health and knowledge management teams at KFL&APH. As the model of care expanded to other PHUs, a new committee was created to ensure a real-time exchange of logistical support. During these meetings, barriers and facilitators to implementing the POC test and treatment are identified, and adaptations were made. These meetings allow project members to ask questions and provide input about POC test and treat implementation and evaluation processes, procurement of tests, and HCP training, and provide updates to the progress of POC test and treat implementation and lessons learned in their respective PHUs.

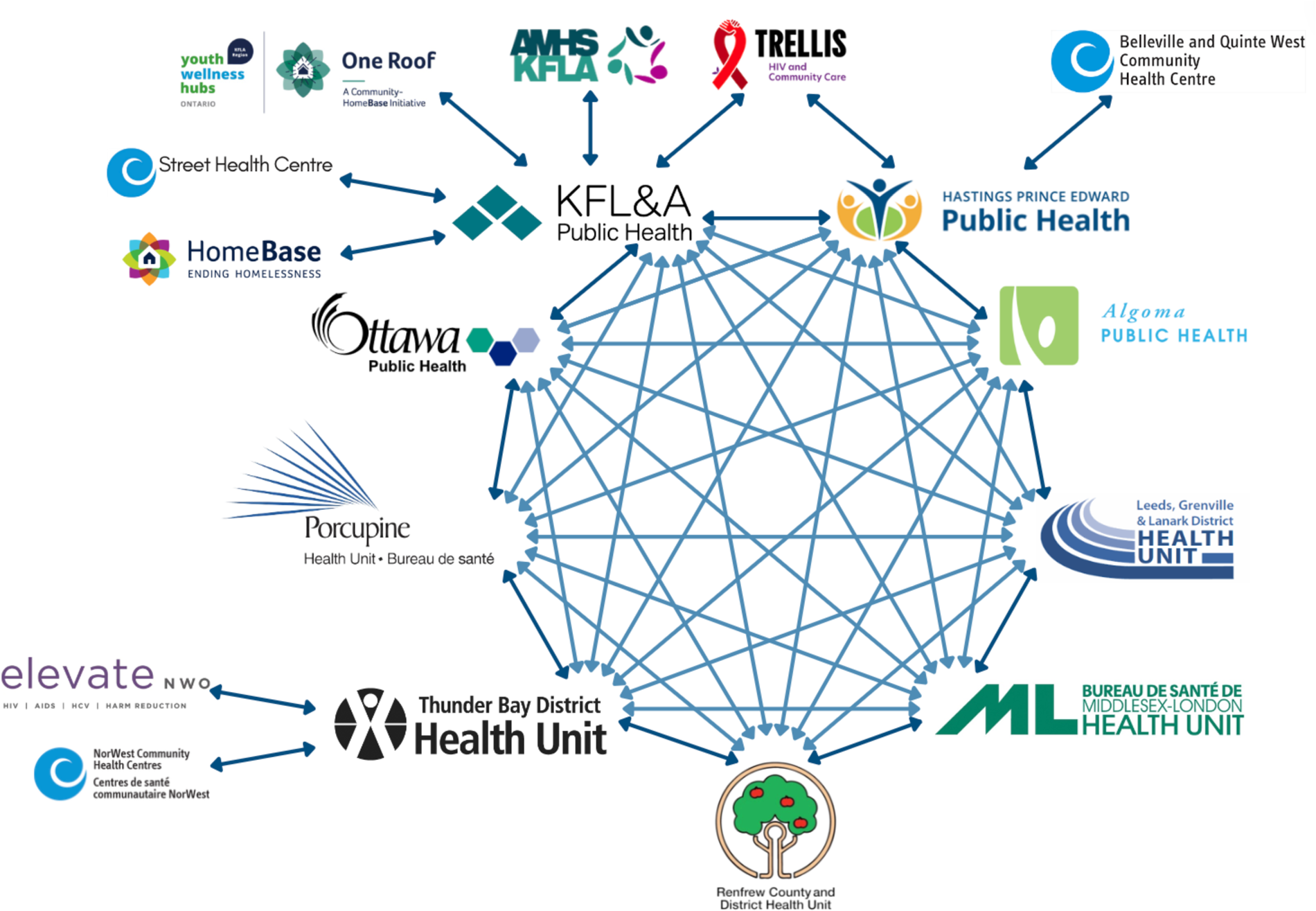

This network will be utilized to create a Community Advisory Group (CAG), which is comprised of individuals representing CBOs and others as identified throughout the process. People with lived experience of homelessness and substance use will be recruited by the CBO advisors to ensure appropriate representation by sex, geography, and risk factors. The CAG will meet monthly to be consulted throughout the research’s planning, data collection, and dissemination to refine objectives and design. Representation from PHO and members of Ontario’s provincial Public Health Sector Coordination Table on syphilis will accelerate knowledge mobilization within and beyond the participating PHUs.

*Analysis/Evaluation:* The REAP Self-Assessment Model, which focuses on Reciprocity, Externalities, Access, and Partnership will be used to evaluate the iKT. Reciprocity will be measured by analyzing the bidirectional flow of knowledge and benefits between researchers and community partners, considering feedback mechanisms, collaborative decision-making processes, and the active involvement of community partners in shaping research directions (53). Externalities will be examined to identify broader societal impacts beyond immediate partners, evaluating increased awareness and positive attitude changes. Access will be assessed for sustained engagement, ensuring ongoing partner access to information and resources. The Partnership dimension will gauge the quality and depth of collaborations, considering engagement levels, communication, and joint decision-making, with collaborative outputs indicating partnership success.

## PATIENT AND PUBLIC INVOLVEMENT

Through the collaborative development of objectives, consultation with a network of PHUs and CBOs, and creating and integrating a CAG, this project engages the community throughout the research process. This protocol was co-developed jointly by KFL&APH and researchers at Queen’s University and integrates the expertise and insights from this multidisciplinary network, including individuals with lived experience of homelessness and/or substance use, following multiple meetings with knowledge users (e.g. PHO, CBOs, and other PHUs). While this protocol outlines our intended objectives and methods, this study is evolving through the input of our CAG.

## ETHICS AND DISSEMINATION

### Dissemination

We will develop a report in lay language outlining the implementation of the research project and key findings, which will be made widely available to interested parties, and develop mini summaries of key findings to be disseminated through the social media accounts of all partnered academic institutions, PHUs, and CBOs. Team members will facilitate webinars to broadly share key findings and lessons learned with KUs and community members. Full research results will be disseminated to all levels of public health (PHUs and PHO) to ensure our results are integrated into practice and with the broader scientific community through peer-reviewed journal articles (open-access) and presentations at relevant Canadian and international conferences. Lastly, we will organize a one-day interdisciplinary forum after the project for our network of CBOs, PHUs, researchers, trainees, and decision-makers from local and provincial PHUs. The purpose is to unite this invested group of individuals to share local community practice and the latest research findings, allowing for continued dialogue and knowledge exchange among our partners.

### Ethics

This study will be conducted in accordance with the Canadian Tri-Council Policy Statement Version 2 and the latest Seoul revision of the Declaration of Helsinki.

As part of routine nursing outreach and best practice, individuals will be verbally informed of the risks and benefits of any testing before any procedures occur. Given the setting and systematic marginalization experienced by the target group, written informed consent to use personal data in an evaluation will not be pursued for this project, as this may dissuade individuals from getting tested altogether. Instead, verbal consent to use data in research or evaluation will be obtained by outreach nurses or other HCPs administering the tests. For the staff surveys, a written letter of information will be provided with the survey, with completion representing consent. Written informed consent will be obtained from study participants for all focus groups and interviews.

Participants of the POC test and treat intervention, PHUs, will have the option to provide five-dollar incentives, usually in the form of a gift card. In certain instances, incentives may not be given depending on locations/events and safety, to provide incentives or not. Staff will not receive compensation for completing the online survey. All participants (staff and members from underserved communities) will be compensated $50 per hour for focus groups and individual interviews in recognition of their time and contribution.

All data and documentation will be managed according to the Personal Health Information Protection Act (PHIPA) and agency policies on safeguarding personal health information. Data will be de-identified by each PHU and sent to KFL&APH via Medallia.

The protocol for the SPRITE study was approved by the Queen’s University Research Ethics Board (REB) at the time of the initial manuscript submission —file numbers 6040037, 6039604, and 6041372. Participant recruitment began in June 2023 and will continue until March 2025 based on current research funding.

## DISCUSSION

A POC test for syphilis is new in Canada. Many public health agencies are interested in how frontline public health nurses can implement POC tests. Underserved populations, including those who are un(der)housed, street-involved, or use injection drugs, are known to be disproportionately affected by syphilis (8, 12, 13). Providing accessible STBBI services to these individuals improves individual-level health outcomes and prevents ongoing transmission. While treatment for syphilis can be completed based on symptom and risk assessment without requiring test results, in outreach contexts, there is often no access to a clinician who has prescribing abilities. Testing yields outcomes that treatment alone cannot, such as disease surveillance and partner notification. The medical directive allowing trained outreach nurses to treat suspected syphilis cases based on POC test results is a key component to making this model of care accessible to underserved communities. While this intervention has theoretical potential, there is currently a knowledge gap on the real-world effectiveness in a community outreach setting and the role it could play in improving the health of individuals of underserved communities, recognizing the multitude of barriers faced by and the competing survival needs of this population. This study looks to address this gap to inform improvements for future implementation, build capacity for other PHUs wanting to implement POC tests and provide information necessary for program decision-making. In conclusion, this study has immense potential to generate high-impact observations, tools, and knowledge that will have long-lasting benefits for public health. Overall, we expect that our results will inform clinical practice and policies around providing STBBI services to underserved populations, thereby improving health outcomes.

## Data Availability

N/A- the study is still in the data collection phase.

## AUTHOR CONTRIBUTIONS

Principal Investigators: SS and MC

Funding Acquisition: SS and MC

Investigation: SS, MC, LM, FM, SZ, KB, JB, MH, KM, JMC, POB, MRG, BS, VT, NS, SV, and GD

Methodology: SS, MC, FM, SZ, and LM

Project Administration: MC, MH, LM and SS

Writing-original draft: LM, SS and MC

Writing-review and editing: SS, MC, LM, MH, POB, NL, FM, SZ, BS, MRG, KM, KB, JB, GC, GD, DD, AF, FK, JMC, VT, NS, and SV

All authors have read and agreed to the published version of the manuscript.

## FUNDING STATEMENT

This work was supported by the Canadian Institutes of Health Research (CIHR) through a Catalyst (SR8 190795), Operating (AS1-192619), and Knowledge Mobilization (EKS 193138) granted to the NPA SS. The team gratefully acknowledges funding received from PHO through the Locally Driven Collaborative Projects program granted to NPA MC. The views expressed in this publication are the views of the project team and do not necessarily reflect those of Public Health Ontario or CIHR.

SPRITE team would also like to thank Public Health Ontario (PHO) for its support of this project.

POB would like to acknowledge funding from the Ontario HIV Treatment Network, Public Health Ontario, Canadian Institutes of Health Research, the National Microbiology Laboratory, and Health Canada.

